# Humoral and cellular response induced by a second booster of an inactivated SARS-CoV-2 vaccine in adults

**DOI:** 10.1101/2022.08.22.22279080

**Authors:** Felipe Melo-González, Constanza Méndez, Hernán F Peñaloza, Bárbara M Schultz, Alejandro Piña-Iturbe, Mariana Ríos, Daniela Moreno-Tapia, Patricia Pereira-Sánchez, Diane Leighton, Claudia Orellana, Consuelo Covarrubias, Nicolás MS Gálvez, Jorge A Soto, Luisa F Duarte, Daniela Rivera-Pérez, Yaneisi Vázquez, Alex Cabrera, Sergio Bustos, Carolina Iturriaga, Marcela Urzua, María S Navarrete, Álvaro Rojas, Rodrigo Fasce, Jorge Fernández, Judith Mora, Eugenio Ramírez, Aracelly Gaete-Argel, Mónica Acevedo, Fernando Valiente-Echeverría, Ricardo Soto-Rifo, Daniela Weiskopf, Alba Grifoni, Alessandro Sette, Gang Zeng, Weining Meng, CoronaVac03CL Study Group, José V González-Aramundiz, Pablo A González, Katia Abarca, Susan M Bueno, Alexis M Kalergis

**Author notes:** **Corresponding author:** Alexis M Kalergis, Pontificia Universidad Católica de Chile. Av. Libertador Bernardo O’Higgins Nº 340, Santiago 8331010, Santiago, Chile. **Alternate corresponding author:** Susan M Bueno, Pontificia Universidad Católica de Chile. Av. Libertador Bernardo O’Higgins Nº 340, Santiago 8331010, Santiago, Chile. These authors contributed equally to this work (Felipe Melo-González, Constanza Méndez, Hernán F Peñaloza, Bárbara M Schultz).

## Abstract

The SARS-CoV-2 Omicron variant has challenged the control of the COVID-19 pandemic even in highly vaccinated countries. While a second booster of mRNA vaccines improved the immunity against SARS-CoV-2, the humoral and cellular responses induced by a second booster of an inactivated SARS-CoV-2 vaccine have not been studied. In the context of a phase 3 clinical study, we report that a second booster of CoronaVac^®^ increased the neutralizing response against the ancestral virus yet showed poor neutralization against the Omicron variant. Additionally, isolated PBMCs displayed equivalent activation of specific CD4^+^ T cells and IFN-γ production when stimulated with a mega-pool of peptides derived from the spike protein of the ancestral virus or the Omicron variant. In conclusion, a second booster dose of CoronaVac^®^ does not improve the neutralizing response against the Omicron variant compared with the first booster dose, yet it helps maintaining a robust spike-specific CD4^+^ T cell response.

## Introduction

The development of vaccines that grant long-lasting protection against SARS-CoV-2 is essential to control the current COVID-19 pandemic. Although several vaccines were developed in record time, three dynamic phenomena have prevented the global control of the COVID-19 pandemic. First, the continuous emergence of SARS-CoV-2 variants of concern (VOCs), such as Omicron (BA.1, BA.2), and its subvariants (BA.2.12.1, BA.4, BA.5) with high transmissibility and immune evasion profiles^1^. Second, the waning of neutralizing antibodies in fully vaccinated subjects^2^. Third, the difficulties to mass producing and globally distributing enough vaccines or implementing affordable and effective vaccination programs.

Several platforms have been used to develop vaccines against SARS-CoV-2^3^. Due to their novelty, mRNA vaccines (BNT162b2 and mRNA-1273) have been the most studied and are highly effective in protecting individuals from symptomatic infection, severe disease, and death^4,5^. Inactivated virus-based vaccine, a more traditional vaccine platform, has also been used to develop a SARS-CoV-2 vaccine. CoronaVac^®^, an inactivated vaccine developed by Sinovac Life Sciences Co., Ltd. (Beijing, China)^6^, has been administered so far in 52 countries^7^, showing a good safety profile in the population^8-10^ and a robust immune protection against severe disease, hospitalization, and death^11^.

During a phase 3 clinical trial in Chile, our group demonstrated that a two-dose vaccination schedule of CoronaVac^®^ induced a strong neutralizing response and T cell activation against SARS-CoV-2 in adults^12^. Further studies from our laboratory determined that fully vaccinated subjects with CoronaVac^®^ showed a strong production of neutralizing antibodies and IFN-γ production in stimulated peripheral blood mononuclear cells (PBMCs) against different VOCs of SARS-CoV-2, such as Alpha, Beta, Gamma, and Delta^13^.

Different studies have reported a reduction of the neutralizing response against SARS-CoV-2 in immunized subjects with BNT162b2 and mRNA1273 vaccines^14-16^. These studies also showed that a booster dose was required to keep an effective neutralizing response against the ancestral SARS-CoV-2 (WT SARS-CoV-2) and circulating variants at that time^14,15,17^.

Consistently with the mentioned studies, our group reported a considerable reduction of the neutralizing response five months after the administration of the second dose of CoronaVac^®^, response that was recovered after the administration of a booster dose of CoronaVac^®18^. Furthermore, the enhanced neutralizing response initially detected against WT SARS-CoV-2 was effective against the Delta variant, but showed reduced neutralization against the Omicron variant^18^, which is currently the most prevalent variant of SARS-CoV-2 worldwide.

Although previous studies have shown that a second booster with BNT162b2 and mRNA1273 vaccines would prevent the decrease of neutralizing antibodies and may offer protection against symptomatic disease, severe disease, hospitalization, and death caused by the Omicron variant^4,19^, the effect of a second booster dose of CoronaVac^®^ in the humoral and cellular response against SARS-CoV-2, with special emphasis on the Omicron variant, remains to be elucidated. In the present report, we study the dynamics of the humoral and cellular immune responses in individuals that received a second booster of CoronaVac^®^ 6 months after the administration of a first booster of the same vaccine. Our data shows that a second booster of CoronaVac^®^ induces a strong production of antibodies with neutralizing capacities against WT SARS-CoV-2, although it has poor activity against the Omicron variant. We also show that a second booster dose of CoronaVac^®^ is required to keep high levels of SARS-CoV-2-specific CD4^+^ T cells in circulation that are reactive against the WT SARS-CoV-2, the Delta, and the Omicron variants.

## Results

### 1. Participants, sampling and experimental applications included in the study

From a total of 2,302 individuals enrolled in the clinical trial CoronaVac03CL (clinicaltrials.gov #NCT04651790) in Chile (November 2020-to current date) ^12,13,18,20,21^, 138 fully vaccinated subjects with CoronaVac^®^ (0-28 schedule) that received two booster doses were initially considered for this study (Fig 1A). After the exclusion of 51 subjects due to SARS-CoV-2 infection during the trial or missing data, longitudinal analyses of the humoral response were performed in up to 87 subjects, whereas the cellular response was studied in a subgroup of 46 subjects (Fig 1A).

**Figure 1.**
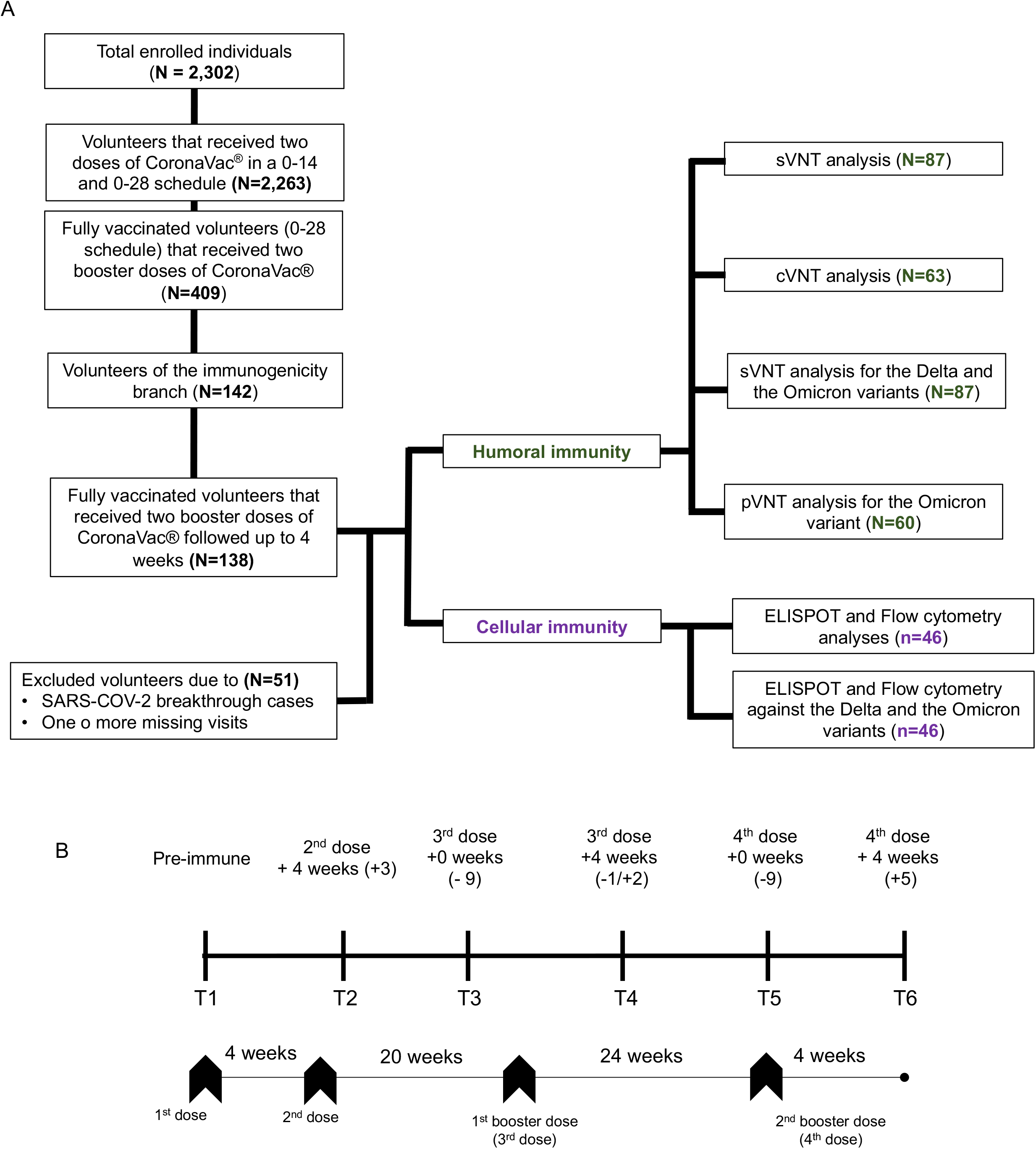
Study profile, vaccination scheme and sampling, enrolled volunteers, and cohort included in the study. (A) Schematic representation and sample distribution of performed experiments. From a total of 138 individuals that received two booster doses of CoronaVac®, the neutralizing antibodies were analyzed in blood samples from 87 volunteers by surrogate virus neutralization test (sVNT), 63 by conventional virus neutralization test (cVNT) and 60 by Pseudovirus-based neutralization assay (pVNT). sVNT and pVNT were used to evaluate the neutralizing response induced by a second booster dose against the Delta and/or the Omicron variants. Cellular immunity was analyzed in blood samples from 46 volunteers at each time point. (B) Blood sampling times, before the first vaccination/pre-immune (T1), at 4 weeks (+3 weeks) after the second dose (T2), before the administration of the third dose (−9 weeks) (T3), at 4 weeks (−1/+2 weeks) after the third dose (T4), before the administration of the fourth dose (−9 weeks) (T5) and at 4 weeks (+5 weeks) after the fourth dose (T6), and vaccination schedule.

Blood samples were collected before vaccination (T1), four to seven weeks after the second dose (T2), at least 9 weeks before the first booster dose (T3), three to six weeks after the first booster dose (T4), at least nine to cero weeks before the second booster dose (T5) and four to nine weeks after second booster dose (T6) (Fig 1B).

### 2. Humoral response against SARS-CoV-2 induced by a second booster dose of CoronaVac^®^

Neutralizing response of serum was evaluated by three different and complementary methodologies, surrogate virus neutralization test (sVNT), conventional virus neutralization test (cVNT), and pseudotype virus neutralization test (pVNT) (see method section). Consistently with previous studies^12,18^, individuals vaccinated with two doses of CoronaVac^®^ presented a significant increase in neutralizing antibodies against WT SARS-CoV-2 four weeks after the administration of the second dose when compared with the pre-immune serum (T1 vs. T2) (16.8 vs. 199.7 GMUs p<0.0001; 2.9 vs. 21.1 GMTs p<0.0001) (Fig. 2A-B, supplementary tables 1 and 3). Then, a significant reduction in the neutralizing antibody against SARS-CoV-2 was observed three to five months (15-20 weeks) after the administration of the second dose when compared with four weeks after the administration of the second dose (T2 vs. T3) (199.7 vs. 53.1 GMUs p<0.0001; 21.1 vs. 10.0 GMTs p<0.0001) (Fig. 2A-B supplementary table 1 and 3). Consistently with other studies^14-16^, the administration of a first booster dose resulted in a rapid improvement of the neutralizing response against WT SARS-CoV-2 after 4 weeks when compared with at least nine weeks before its administration (T3 vs. T4) (53.1 vs. 586.0 GMUs p<0.0001; 10.0 vs. 95.1 GMTs p<0.0001) (Fig. 2A-B supplementary table 1 and 3). Interestingly, the neutralizing capacity against WT SARS-CoV-2 observed six months (24 weeks) after the administration of the first booster dose was partially reduced in comparison to the response detected 4 weeks after the administration of the first booster dose (T5 vs. T4) (220.4 vs. 586.0 GMUs p<0.001; 54.9 vs. 95.1GMTs p>0.05) (Fig. 2A-B, supplementary table 1 and 3). Finally, 4 to 9 weeks after the administration of a second booster dose, an increase in the neutralizing response against WT SARS-CoV-2 was observed when compared with the time before the administration of the second booster dose (T5 vs. T6) (220.4 vs. 549.2 GMUs p<0.001; 54.9 vs. 149.3 GMTs p>0.05) (Fig. 2A-B, supplementary table 1 and 3). Importantly, the neutralization response against WT SARS-CoV-2 observed after the administration of the second booster dose was not significantly higher than the response induced by the first booster dose (T4 vs. T6) (586.0 vs. 549.2 GMUs p>0.05; 95.1 vs. 149.3 GMTs p>0.05) (Fig. 2A-B, supplementary table 1 and 3). This data indicates that a second booster dose of CoronaVac^®^ is required to keep high levels of neutralizing antibodies against WT SARS-CoV-2. Moreover, given that the neutralization observed after the first and the second booster dose was equivalent, we hypothesize that the neutralizing response against SARS-CoV-2 induced by the administration of the first booster dose of CoronaVac^®^ has reached a peak of neutralizing antibodies that are sustained by the administration of a second booster dose.

**Figure 2.**
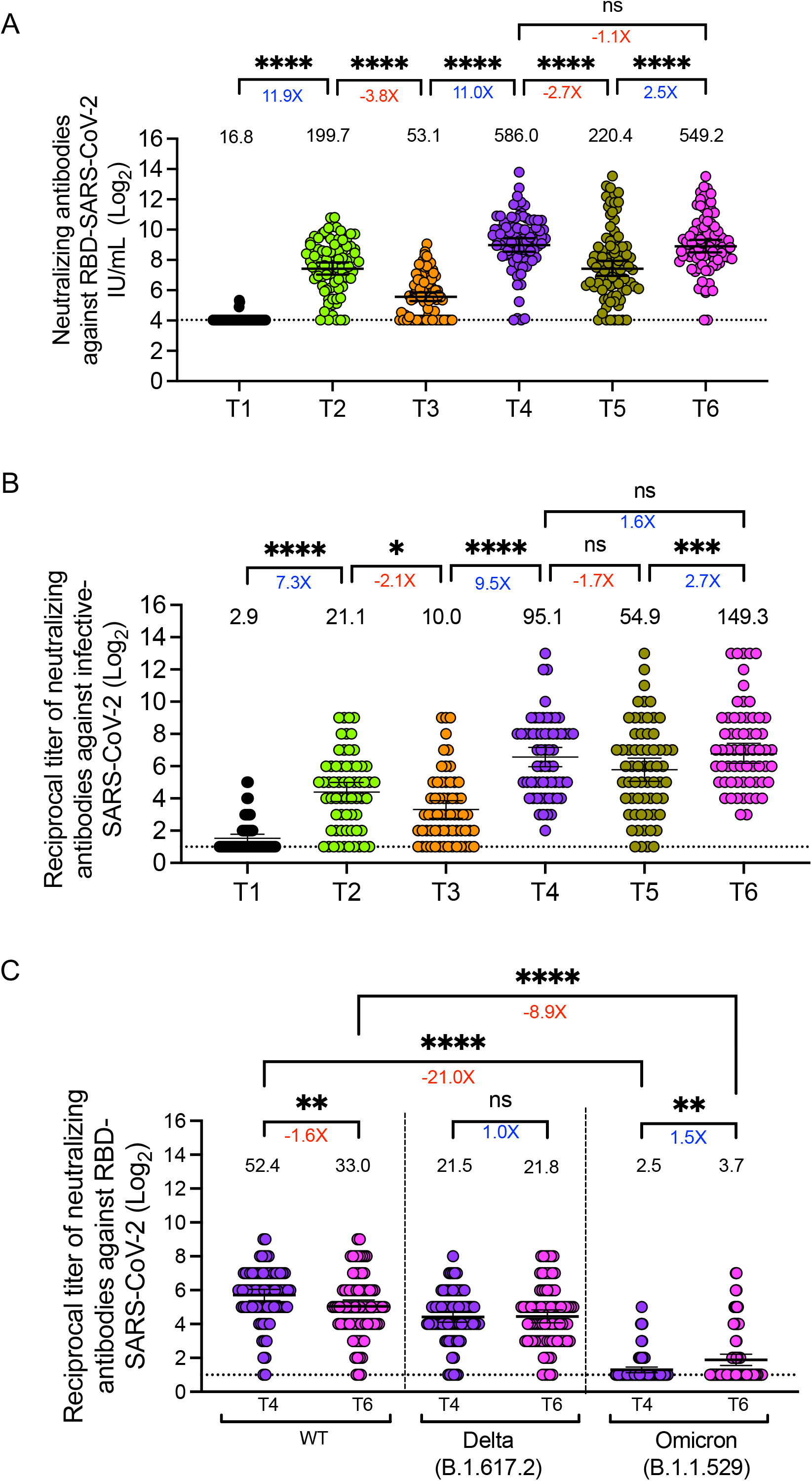
Humoral response of volunteers who received a second booster dose of CoronaVac^®^. The neutralizing capacity of circulating antibodies in adults was evaluated in Blood samples collected before the first vaccination/pre-immune (T1), at 4 weeks (+3 weeks) after the second dose (T2), before the administration of the third dose (−9 weeks) (T3), at 4 weeks (−1/+2 weeks) after the third dose (T4), before the administration of the fourth dose (−9 weeks) (T5) and at 4 weeks (+5 weeks) after the fourth dose (T6), and vaccination schedule. (A) Neutralizing capacity of circulating antibodies against SARS-CoV-2 in serum of 87 volunteers was determined by a surrogate Viral Neutralization Test (sVNT) expressed as IU/ml. (B) Reciprocal dilution of sera required to prevent in vitro infection obtained sera from 63 adults required to prevent in vitro infection of Hela Cells. Numbers on top of each data set represents the GMT, and horizontal lines represent the 95% CI. (C) Geometric mean titter (GMTs) of neutralizing antibodies against the WT-spike, Delta-spike and Omicron-spike proteins detected in the serum of 87 volunteers immunized with CoronaVac® through sVNT. Dashed line: limit of detection. Red values under the significance line: indicate a decrease in the means of the two compared time points; Blue values: indicate an increase in the means of the two compared time points. (A-B) Data was analyzed with ANOVA with the Geisser-Greenhouse correction test followed by a post-hoc Sidak multiple test. *P<0.05; **P<0.01; ***P<0.001; ****P<0.0001. (C) Two-way ANOVA test followed by a post-hoc Dunn’s multiple test. *P<0.05; **P<0.01; ***P<0.001, ****P<0.0001.

The administration of the second booster dose of CoronaVac^®^ kept the seropositivity rate higher than 96% for both sVNT and cVNT, with seroconversion levels of 93.1% and 95.2% for sVNT and cVNT, respectively (supplementary table 3).

Next, we evaluated whether the neutralizing antibodies generated after the second booster dose was effective against the Delta (B.1.617.2) and the Omicron (B.1.1.529) variants. Although serum from vaccinated individuals collected four weeks after the second booster dose presented a mildly reduced ability to neutralize WT SARS-CoV-2 when compared with samples collected four weeks after the first booster dose (Fig 2C), the neutralization response against WT SARS-CoV-2 was significantly higher when compared with the Delta (B.1.617.2) and especially with the Omicron (B.1.1.529) variants (33.0 vs. 3.7 GMTs p<0.001, 8.9-fold reduction) (Fig 2C, supplementary table 2). Moreover, as compared with the first booster dose, the administration of the second booster dose of CoronaVac^®^ did not significantly impact the seropositivity rate (93.1% vs. 94.3%) and the seroconversion rate (80.5% vs. 67.8%) against the Delta variant, although it slightly increased the seropositivity rate (17.2% vs. 32.2%) and the seroconversion rate (4.6% vs. 18.4%) against the Omicron variant (supplementary table 4).

Neutralization against the Omicron variant was also confirmed with a pseudotype-based neutralization assay in sixty subjects (supplementary figure 1) and indicate that a second booster dose of CoronaVac^®^ maintains high neutralizing antibody levels against WT SARS-CoV-2 that reduced neutralization capacity against the Omicron variant.

### 3. Cellular response against SARS-CoV-2 induced by a second booster dose of CoronaVac®

We next evaluated the cellular response in a subgroup of fully vaccinated volunteers (n=46) that received two booster doses of CoronaVac^®^ (Fig 1A). PBMCs were stimulated with mega-pools of peptides of SARS-CoV-2 theoretically able to activate CD4^+^ T cells (S+R) and CD8^+^ T cells (CD8A+B)^22^ and SARS-CoV-2-specific OX40^+^CD137^+^CD4^+^T cells (AIM^+^CD4^+^ T cells), as well as SARS-CoV-2 specific CD69^+^CD137^+^CD8^+^ T cells (AIM^+^CD8^+^ T cells) were quantified by flow cytometry. Our data show that the activation of SARS-CoV-2-specific AIM^+^CD4^+^ T cells was higher after the second dose of CoronaVac^®^ in comparison with the pre-immune sample (0.13% vs. 0.33% p=0.0241) (Fig. 3A; supplementary table 5). Although no significant increase in the percentage of AIM^+^CD4^+^ T cells was observed after the administration of the first or the second booster dose, the activation of AIM^+^CD4^+^ T cells remained stable over time, showing only a significant decrease 4-6 months after the first booster dose, that is successfully recovered after the administration of the second booster dose (Fig 3A, supplementary table 5). Further, we did not detect a significant activation of SARS-CoV-2-specific AIM^+^CD8^+^ T cells in fully vaccinated volunteers with CoronaVac^®^ after the administration of the first or the second booster dose (Fig. 3B; supplementary table 5).

**Figure 3.**
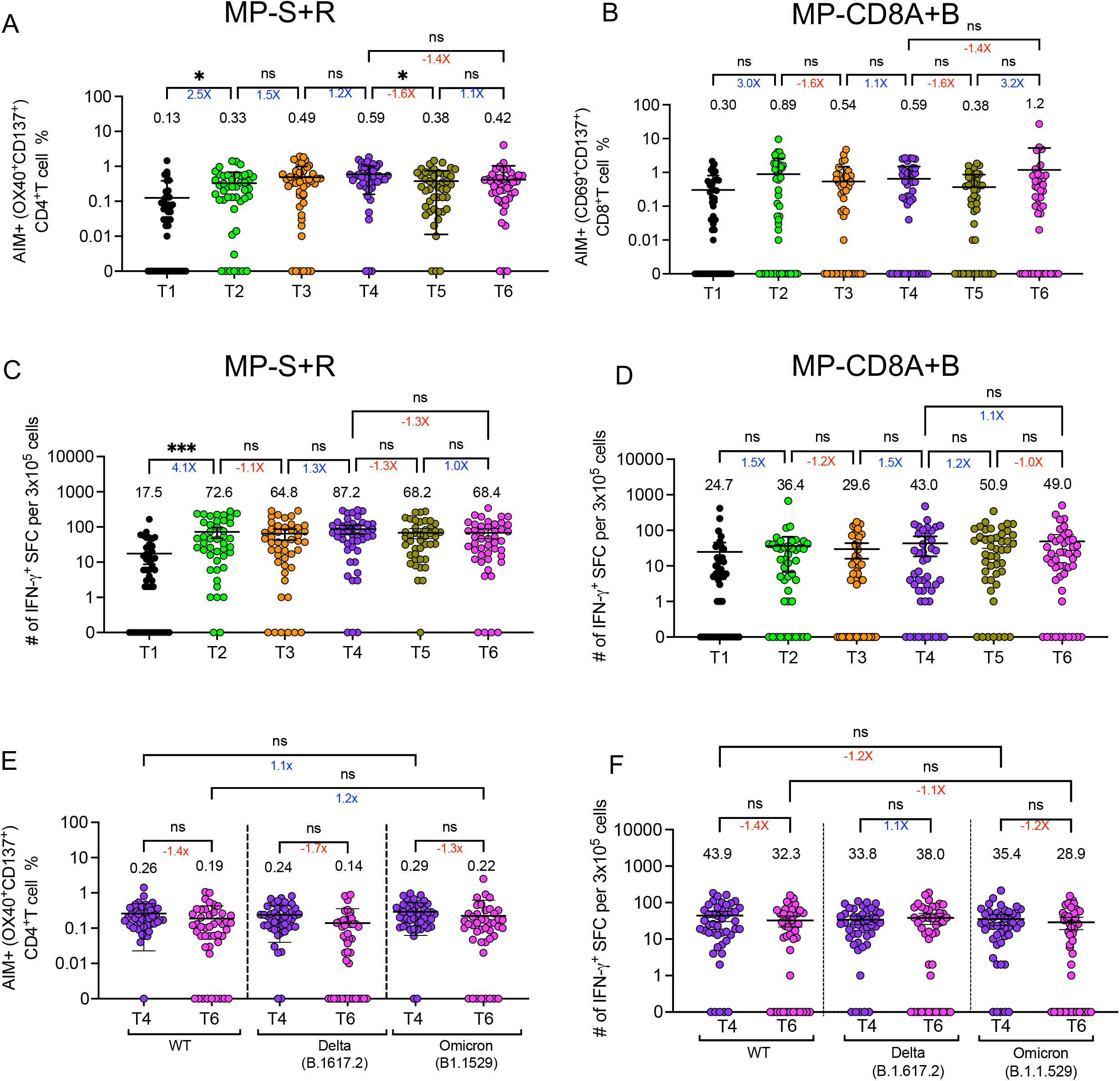
Cellular response of volunteers that received a second booster dose of CoronaVac^®^. Cellular response in adults was evaluated in PBMCs collected before the first vaccination/pre-immune (T1), at 4 weeks (+3 weeks) after the second dose (T2), before the administration of the third dose (−9 weeks) (T3), at 4 weeks (−1/+2 weeks) after the third dose (T4), before the administration of the fourth dose (−9 weeks) (T5) and at 4 weeks (+5 weeks) after the fourth dose (T6), and vaccination schedule. (A) The percentage of AIM+ (OX40+CD137+) CD4+ T cells and (B) AIM+ (CD69+CD137+) CD8+ T cells was determined in PBMCs of 46 adult volunteers by flow cytometry. PBMCs were stimulated for 24h with mega-pools of peptides derived from proteins of WT SARS-CoV-2. The number of IFN-g producing SFCs was determined by ELISPOT upon stimulation for 48h with mega-pools of (C) S+R peptides or with (D) CD8A+B peptides. (E) The percentage of AIM+ (OX40+CD137+) CD4+ T cells from PBMCs of 46 adult volunteers were analyzed by flow cytometry after the stimulation for 24h with mega-pools of peptides derived from the Spike protein of the WT SARS-CoV-2, the Delta and the Omicron variants. (F) The number of IFN-g producing SFCs was determined by ELISPOT assays PBMCs of 46 adult volunteers were analyzed by flow cytometry after the stimulation for 48h with mega-pools of peptides derived from the Spike protein of the WT SARS-CoV-2, the Delta and the Omicron variants. Horizontal lines represent mean and 95% CI. Flow cytometry data was normalized against the DMSO control. (A-D) Data was analyzed using a non-parametric Friedman test followed by a post-hoc Dunn’s test for multiple comparisons. *P<0.05; **P<0.01; ***P<0.001; ****P<0.0001. (E-F) Two-way ANOVA test followed by a post-hoc Dunn’s multiple test. *P<0.05; **P<0.01; ***P<0.001, ****P<0.0001.

Next, we evaluated the production of IFN-γ in stimulated-PBMCs by ELISPOT (Fig. 3C-D, supplementary table 5). The production of IFN-γ by stimulated PBMCs was consistent with the activation of CD4^+^ and CD8^+^ T cells in subjects that received a second booster dose of CoronaVac^®^. A significant increase in IFN-γ^+^ SFCs was observed in PBMCs stimulated with MP-S+R after the second dose of CoronaVac^®^ (4.1-fold increase in PBMCs stimulated with MP-S+R p=0.0004) (Fig 3C, supplementary table 5). Moreover, the increase in IFN-γ^+^SFCs observed after the second dose of CoronaVac^®^ remained stable over time and was also observed after the administration of the first and the second booster dose (Fig 3C, supplementary table 5). In contrast, no significant changes in IFN-γ^+^SFCs were observed in PBMCs stimulated with MP-CD8A+B after the second dose of CoronaVac^®^ (1.5-fold increase in PBMCs stimulated with MP-CD8A+B p>0.05) (Fig. 3D) nor after the administration of a first or second booster of CoronaVac^®^ (Fig 3D, supplementary table 5).

Even though we did not detect a significant increase in the frequency of AIM^+^CD4^+^T cells after the administration of the first or the second booster doses of CoronaVac^®^, we observed important changes in the frequency of volunteers with detectable levels of AIM^+^CD4^+^T cells over time. Whereas 13.0% (6/46) of volunteers presented detectable levels of SARS-CoV-2-specific AIM^+^CD4^+^T cells before vaccination, two doses of CoronaVac^®^ increased this frequency to 34.8% (16/46), frequency that was further increased after the first booster of CoronaVac^®^, where 65.2% (30/46) of subjects showed detectable levels of SARS-CoV-2-specific AIM^+^CD4^+^T cells (Table 1). Consistently, the frequency of subjects whose PBMCs produced IFN-γ after stimulation with MP-S+R increased after the administration of CoronaVac^®^, starting from 17.4% (8/46) before vaccination to 47.8% (22/46) after two doses of CoronaVac^®^ and to 67.4% (31/46) after the first booster of CoronaVac^®^ (Table 1). Interestingly, the second booster of CoronaVac^®^ (45.7%, 21/46) did not increase the frequency of volunteers that have specific SARS-CoV-2 specific AIM^+^CD4^+^T cells in circulation as compared with the first booster dose, although it seems that it is required to prevent the decrease in frequency of volunteers with SARS-CoV-2 specific AIM^+^CD4^+^T cells observed right before the administration of the second booster dose (41.3%, 19/46) (Table 1). Moreover, the second booster of CoronaVac^®^ did not significantly affect the progressive decrease of IFN-γ production by stimulated PMBCs (50% 23/46) in comparison with the response induced by the first booster dose, where 67.4% of the volunteers (31/46) showed production IFN-γ production (Table 1).

**Table 1:**
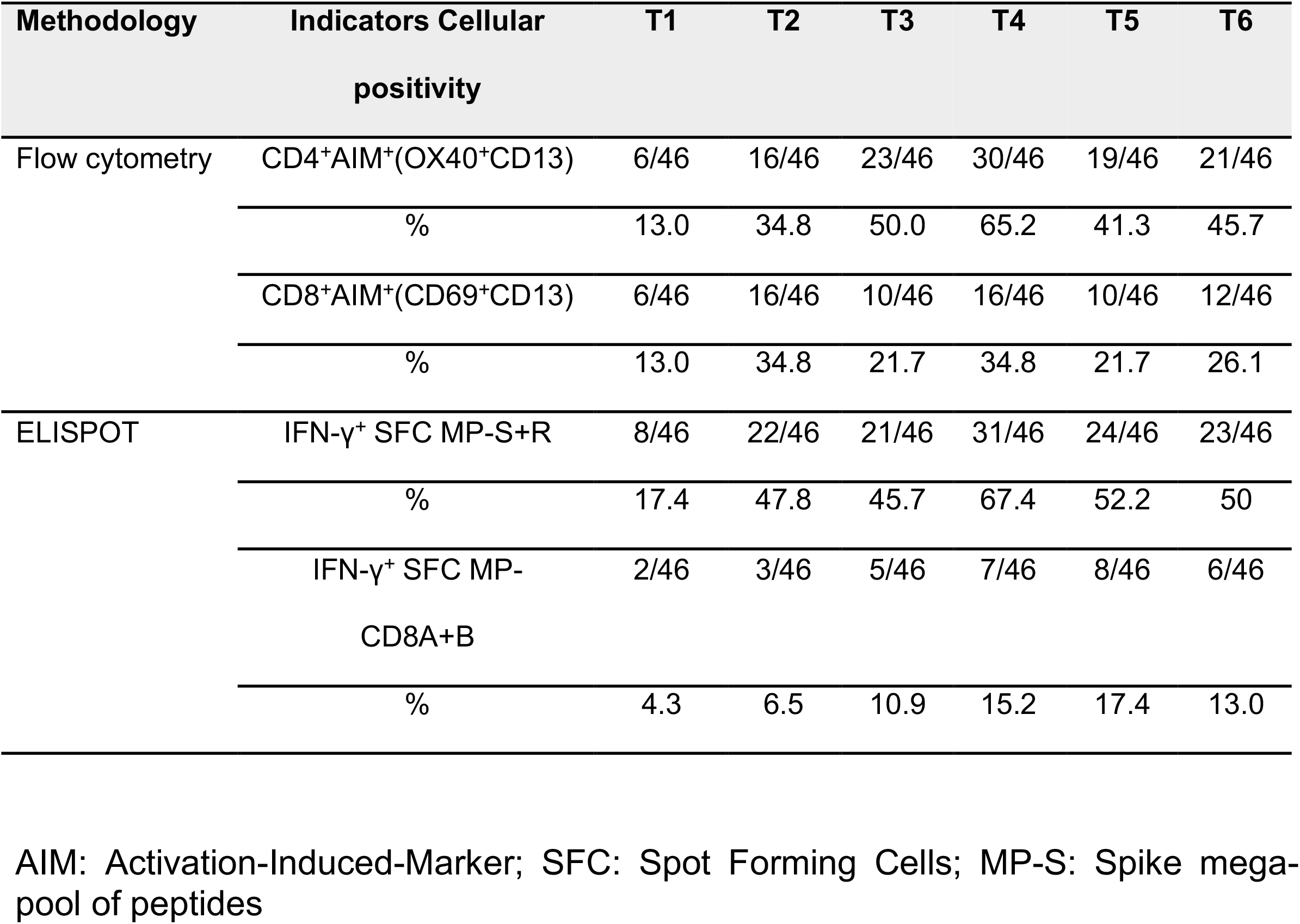
Frequency of volunteers with detectable T cell response against WT SARS-CoV-2.

A mild increase in the frequency of volunteers that showed AIM^+^CD8^+^ T cells after two doses of CoronaVac^®^ (34.8%, 16/46) was observed as compared to before vaccination (13.0%, 6/46) (Table 1). Even though the administration of a first booster of CoronaVac^®^ (34.8%, 16/46) was necessary to keep the frequency of volunteers with detectable levels of SARS-CoV-2-specific AIM^+^CD8^+^ T cells, the second booster dose (26.1%, 12/46) did not prevent the reduction of the positivity of SARS-CoV-2-specific AIM^+^CD8^+^ T cells in our cohort (Table 1). In addition, when PBMCs were stimulated with MP-CD8A+B, the frequency of volunteers that showed IFN-γ production reached a peak of 17.4% (8/46) right before the administration of a second booster that slightly decreased after the administration of the second booster dose of CoronaVac^®^ (13.0%, 6/46) (Table 1).

Finally, we evaluated the activation of AIM^+^CD4^+^ T cells and IFN-γ^+^ production by PBMCs in response to a mega-pool of peptides derived from the Spike protein of the Delta and the Omicron variants by flow cytometry and ELISPOT, respectively, four weeks after the administration of the first booster dose and four weeks after the administration of the second booster dose. Our data show that the activation AIM^+^CD4^+^ T cells and IFN-γ^+^SFCs were equivalent when PBMCs were stimulated with a mega-pool of peptides derived from the WT SARS-CoV-2, the Delta, or the Omicron variants (Fig. 3E-F) and no major differences were detected in the activation of AIM^+^CD4^+^ T cells and IFN-γ^+^SFCs in PBMCs, between the first and the second booster dose against each of these variants (Fig. 3E-F). Importantly, we detected a small decrease in the percentage of subjects that presented Spike-specific CD4^+^AIM^+^ against the WT, Delta and Omicron variants after the second booster compared with after the first booster dose (supplementary table 6). Importantly, the frequency of volunteers whose PBMCs produced IFN-γ remained constant after the stimulation of MP-S from WT and the Delta variant and only showed a mild decreased after MP-S from the Omicron variant (supplementary table 6).

These data show that CoronaVac^®^ induces a robust CD4^+^ T cells response able to react against the Delta and the Omicron variants that remains high after a second booster dose even though it showed a progressive decrease in its positivity. Although the humoral immunity against Delta and Omicron variants decreases, cellular immunity remains robust across time and reacts against these variants.

## Discussion

In line with other reports^4,19^, our data show that a second booster dose of CoronaVac^®^ restores the neutralizing response against the RBD of SARS-CoV-2 to similar levels reached after the administration of the first booster dose.

Neutralizing antibodies induced by vaccines has been acknowledged as the first line of defense against SARS-CoV-2 infection. However, follow-up studies have shown that fully vaccinated individuals show a gradual decrease in their levels of circulating neutralizing antibodies over time^14-16,18^. This response can be restored with the administration of a booster dose^14,18^, and some studies have even shown that a second booster dose may grant better protection against severe disease, hospitalization, and death due to COVID-19^4,23^.

Our data show that despite a second booster dose induces the production of neutralizing antibodies against WT SARS-CoV-2, these antibodies only weakly neutralized the Omicron variant. These results are consistent with other studies that show that the Omicron variant and its subvariants are incredibly efficient in evading serum neutralization from individuals who have received one or two booster doses^1,19,24^. Therefore, it is unlikely that nowadays, the increased protection against severe disease, hospitalization, and death granted by a second booster dose could be mainly mediated by the action of neutralizing antibodies.

Nonetheless, we detected a slight but non-significant increment of 1.6-fold change in the neutralization of WT SARS-CoV-2 between four weeks after the first booster dose and four weeks after the second booster dose by cVNT (Fig 1B). This modest increase in WT SARS-CoV-2 neutralization granted by the second booster was not observed through sVNT (Fig 1A, C). sVNT directly neutralizes the binding of the RBD with ACE-2, neglecting the potential role of neutralizing antibodies against other portions of the Spike protein and even against other viral proteins. Therefore, it is possible that neutralizing antibodies against other domains of the Spike protein and potentially against other viral proteins such as the membrane or the nucleocapsid proteins not considered in the sVNT approach may actively participate in viral neutralization.

The second layer of protection corresponds to the proliferation, activation, and activity of T cells. Currently, the leading hypothesis regarding the protective mechanism of SARS-CoV-2 vaccines against severe disease and death relies on the induction of long-lasting T cells responses rather than on the availability of circulating neutralizing antibodies^25,26^.

In the present study, we report that the activation of AIM^+^CD4^+^ T cells in fully vaccinated individuals remains detectable over time after administering a first and a second booster of CoronaVac^®^. In addition, one study has identified a robust activation of CD4^+^OX40^+^CD137^+^ T cells, CD4^+^OX40^+^sCD40L^+^ T cells, and follicular CD4^+^CXCR5^+^OX40^+^ T cells in individuals that received two doses of mRNA-1273, BNT162b2 or NVX-CoV2373 (Novavax), and one dose of Ad26.COV2.S (Janssen)^27^. Our data is consistent with these findings, although the ability of CoronaVac^®^ to induce a robust memory T cell response remains to be elucidated. On the other hand, it has been reported a significant expansion and activation of IFN-γ^+^CD8^+^ T cells in PBMCs of individuals vaccinated with two doses of mRNA-1273, BNT162b2, and NVX-CoV2373, and one dose of Ad26.COV2.S^27^. However, we did not find a significant increase in CD8^+^AIM^+^ T cells (Fig. 3B) nor a significant increase in IFN-γ production (Fig 3D) in stimulated PBMCs from subjects immunized with CoronaVac^®^ when compared with the pre-immune sample. In contrast, previous studies have described that two doses of CoronaVac^®^ induce the production of IFN-γ by CD8^+^ T cells^28,29^. Therefore, more exhaustive studies aimed to understand how CoronaVac^®^ influences the expansion and activation of CD8^+^ T cells, memory T cells, and memory B cells are required to fully elucidate the protective mechanisms driven by CoronaVac^®^ in immunized adults, children, and the elderly.

Four interesting findings emerge from our study. First is the presence of neutralizing antibodies (Fig 2A-B) in pre-immune samples of some individuals. Most of the pre-immune neutralizing response was identified against the infective virus. Asymptomatic SARS-CoV-2 infections^30^ and antibodies generated against non-spike proteins of seasonal coronaviruses^31^ could explain this response, although serologic studies are needed to confirm these hypotheses. Moreover, our data show that whereas 6/46 of the analyzed volunteers presented SARS-CoV-2-specific AIM^+^CD4^+^ or AIM^+^CD8^+^ T cells before the administration of CoronaVac^®^; 25/46 and 34/46 volunteers did not present AIM^+^CD4^+^ or AIM^+^CD8^+^ T cells, respectively after the second booster of CoronaVac^®^. A previous study has shown a possible cross-reactive response of seasonal coronaviruses-specific T cells against SARS-CoV-2^22^, and it is likely the reason why some volunteers presented SARS-CoV-2 specific T cells before vaccination, although asymptomatic infections cannot be ruled out. Further studies are needed to fully understand the factors involved in the generation and maintain of an efficient T cell response and the effect on vaccine-induced protection.

Finally, our data show that the effect on humoral and cellular responses of the second booster dose, is different when compared with the effect of the first booster dose. Whereas our data and several other studies have shown that a first booster increase the humoral and cellular response against SARS-CoV-2^14,15,17,18^, the administration of a second booster dose seems to maintain the global neutralizing response, the activation of AIM^+^CD4^+^ T cells and IFN-γ by stimulated PBMCs against WT SARS-CoV-2 and the Omicron variant reached by the first booster dose. Importantly, in terms of the percentage of individuals able to respond against the virus, our data show that even though the second booster restores seropositivity and seroconversion to equivalent levels observed after the first booster dose, it seems to be insufficient to restore the frequency of volunteers with detectable SARS-CoV-2-specific AIM^+^CD4^+^ T cells and IFN-γ by stimulated PBMCs to levels observed after the first booster dose.

Controlling the COVID-19 pandemic requires multiple efforts to prevent severe disease and death of infected patients and reduce viral infection and circulation in the community.

The data provided in this report and other studies suggest that CoronaVac^®^ and other current vaccines effectively protect the population from severe disease, hospitalization, and death. However, the immune response induced by these vaccines poorly neutralizes the circulating SARS-CoV-2 variants and cannot prevent viral infection.

Therefore, new strategies that include the design of new vaccines that target the current variants, new types of vaccines that enforce immunity in the upper respiratory tract, and global vaccine distribution programs are essential to control and end the COVID-19 pandemic.

## Methods

### Volunteers and sample collection

Blood samples were obtained from volunteers recruited in the clinical trial CoronaVac03CL (clinicaltrials.gov #NCT04651790) in Chile (November 2020-to current date)^12,13,18,20,21^.

Of the 2,302 individuals enrolled at baseline, 409 subjects entered the study and received two doses with the homologous CoronaVac^®^ 0-28 schedule, and 138 subjects ended up receiving two booster doses of CoronaVac^®^. In addition, blood samples from 87 subjects were collected from before vaccination (T0) to up to four-nine weeks after the second booster administration (T6). Volunteers who did not have a blood sample at some of the planned times of the study or who had previously exceeded the definitive time limit and volunteers who had developed SARS-COV-2 during the study were excluded (Figure 1A).

Volunteers received two doses of CoronaVac® (3 μg or 600SU of inactivated SARS-CoV-2 inactivated in the presence of alum adjuvant) in a four-week interval (0-28 days), a booster dose five months after the second dose, and a second booster dose 6 months after the first (Figure 1B). Blood samples collected 6 times were analyzed: T1: Pre-immune, T2: 2^nd^ dose + 4 weeks (+3 weeks), T3: before the administration of the third dose (−9 weeks), T4: at 4 weeks (±2 weeks) after the third dose, T5: before the administration of the fourth dose (−9 weeks), T6: and at 4 weeks (+5 weeks) after the fourth dose (T6), and vaccination schedule.

### Experimental procedures

Neutralizing antibodies against RBD of WT SARS-CoV-2 were measured in the serum of 87 volunteers recruited in the clinical trial CoronaVac03CL (clinicaltrials.gov #NCT04651790). Briefly, blood samples were obtained at T1, T2, T3, T4, T5, and T6. Neutralization of SARS-CoV-2 RBD by circulating antibodies at each time point was evaluated by a surrogate virus neutralization test (sVNT) (Genscript Cat#L00847-A)^12^. The antibody levels in international units per mL (IU/mL) were estimated by interpolating the sVNT absorbance data in the standard curve made with the WHO International Standard 20/136, using the 4-parameter Logistic model. In addition, sVNT was used to evaluate the neutralizing capacity of the tools against the Delta and Omicron variants, the RBDs for the S protein of SARS-CoV-2 for Delta (Cat# Z03516) and Omicron B.1.1.529 (Cat#Z03730) in 87 volunteers.

Infective virus neutralization assays were performed as previously described^12,18^. Briefly, Vero E6 cells (4×10^4^ cells/well) were plated in 96-well plates. 100 µL of 33782CL-SARS-CoV-2 (100 TCID50) were incubated with serial dilutions of heat-inactivated individual serum samples (dilutions of 1:4, 1:8, 1:16, 1:32, 1:64, 1:128, 1:256, and 1:512) for 1h at 37 °C. Then, the mix was added to the 96-well plates with the Vero E6 cells, and the cytopathic effect was analyzed after 7 days. A serum sample from uninfected patients (negative control) and a neutralizing COVID-19 patient serum sample (positive control) was used for each test.

A pseudotyped virus neutralization test (pVNT) assay was performed to assess the capacity of the antibodies against SARS-CoV-2 VOC in samples from forty-eight volunteers as previously reported^32^. Briefly, a HIV-1 backbone expressing firefly luciferase as a reporter gene and pseudotyped with the SARS-CoV-2 spike glycoproteins (HIV-1-SΔ19) from lineage B.1 (D614G) and variant Omicron (A67V, ΔH69-V70, T95I, Y145D, ΔG142 -V143-Y144, ΔN211, EPE 213-214, G339D, S371L, S373P, S375F, K417N, N440K, G446S, S477N, T478K, E484A, Q493R, G496S, Q498R, N501Y, T547K, D614G, H655Y, N679K, P681H, N764K, N865K, Q954H, N969K, L981F) was prepared. Serum samples were diluted, and the estimation of the ID80 was obtained using a 4-parameter nonlinear regression curve fit measured as the percent of neutralization determined by the difference in average relative light units (RLU) between test samples and pseudotyped virus controls.

Seropositivity is considered when titers are increased compared to pre-immune condition^33^. On the other hand, seroconversion was thought to be when the titer of neutralizing results increased 4 times with respect to the pre-immune condition.

The expression of Activation-Induced Markers (AIM) by T cells and the number of Spot Forming Cells (SFC) for IFN-γ were determined by ELISPOT and were evaluated by flow cytometry in a subgroup of 46 volunteers as previously described^12,13,18^. Briefly, PBMCs were stimulated with mega-pools (MPs) of peptides derived from SARS-CoV-2: MP-S, MP-R, MP-CD8-A, MP-CD8-B, Delta, and Omicron for 24h or 48h for flow cytometry and ELISPOT respectively. Then, 3-5 × 10^5^ cells were plated and stimulated with MP-S, MP-R, MP-CD8A, MP-CD8B, DMSO (negative control), or Phorbol-12-acetate (1.62mM) (Sigma, #P8139)/Ionomycin (0.6mM) (Sigma #I0634)^22^. Antibodies used to identify AIM^+^CD4^+,^ and AIM^+^CD8^+^ T cells by flow cytometry are detailed in supplementary table 7. Samples were analyzed in a BD LSR-FORTESSA flow cytometer located in the flow cytometry core at the Pontificia Universidad Católica de Chile. For AIM^+^CD4^+,^ and AIM^+^CD8^+^ T cells analyses, the percentage of DMSO was subtracted from each stimulated sample to subtract the background. The positivity threshold for AIM^+^CD4^+^ T cell stimulated with MP-S (0.174%) and MP-S+R (0.36%), as well as AIM^+^CD8^+^ T cell stimulated with MP-CD8A+B (0.66%) was calculated using the median twofold standard deviation of each sample of the pre-immune group^22,34^.

For ELISPOT, IFN-γ was measured using a Immunospot^®^ (#hIFNgIL-4M-10) following the manufacturer’s instructions. Plates were read in an Immunospot S6 Micro Analyzer. The positivity threshold for PBMCs stimulated with MP-S (19.31 #SFC) MP-S+R (41.48 #SFC), as well as PBMCs stimulated with MP-CD8A+B (104.54 #SFC) was calculated using the median twofold standard deviation of each sample of the pre-immune group.

#### Ethical considerations

The current study protocol was reviewed and approved by the Institutional Scientific Ethical Committee of Health Sciences at the Pontificia Universidad Católica de Chile (#200708006) and the trial was approved by the Chilean Public Health Institute (#24204/20) and conducted according to the current Tripartite Guidelines for Good Clinical Practices, the Declaration of Helsinki, and local regulations. Informed consent was obtained from all volunteers upon enrollment.

#### Statistical analyses

To statistically compare the neutralizing response against WT SARS-CoV-2, ANOVA with the Geisser-Greenhouse correction test followed by Sidak multiple tests were carried out on the log transformed data. Percentage of AIM^+^CD4^+^ and AIM^+^CD8^+^ T cells and IFN-γ production by stimulated PBMCs against WT SARS-CoV-2 were compared with a non-parametric Friedman test followed by a Dunn’s test for multiple comparisons. To compare the neutralizing and the cellular responses against WT, Delta and Omicron, a two-way ANOVA test followed by a Dunn’s test for multiple comparisons was used.

The significance level was set at 0.05 for all the analyses. All data were analyzed with GraphPad Prism 9.3.1.

## Data Availability

All data produced in the present work are contained in the manuscript

## Funding

The CoronaVac03CL Study was funded by The Ministry of Health, Government of Chile, the Confederation of Production and Commerce (CPC), Chile and SINOVAC Biotech. NIH NIAID, under Contract 75N93021C00016, supports AS and Contract 75N9301900065 supports AS, AG and DW. The Millennium Institute on Immunology and Immunotherapy, **Agencia Nacional de Investigación y Desarrollo (ANID) – Millennium Science Initiative Program – ICN09_016 / ICN 2021_045: Millennium Institute on Immunology and Immunotherapy (ICN09_016 / ICN 2021_045; former P09/016-F)** supports SMB, KA, PAG and AMK; The Innovation Fund for Competitiveness FIC-R 2017 (BIP Code: 30488811-0) supports SMB, PAG and AMK.

## Competing interests

GZ and WM are SINOVAC Biotech employees and contributed to the conceptualization of the study (clinical protocol and eCRF design) and did not participate in the analysis or interpretation of the data presented in the manuscript. A.S. is a consultant for Gritstone Bio, Flow Pharma, ImmunoScape, Moderna, AstraZeneca, Avalia, Fortress, Repertoire, Gilead, Gerson Lehrman Group, RiverVest, MedaCorp, and Guggenheim. La Jolla Institute for Immunology (LJI) has filed for patent protection for various aspects of T cell epitope and vaccine design work. All other authors declare no conflict of interest.

## Acknowledgments

We would like to thank the support of the Ministry of Health, Government of Chile; Ministry of Science, Technology, Knowledge, and Innovation, Government of Chile; The Ministry of Foreign Affairs, Government of Chile, and the Chilean Public Health Institute (ISP). We also would like to thank Rami Scharf, Jessica White, Jorge Flores and Miren Iturriza-Gomara from PATH for their active support in experimental design and scientific discussion. We also thank the Vice Presidency of Research (VRI), the Direction of Technology Transfer and Development (DTD) and the Legal Affairs Department (DAJ) of the Pontificia Universidad Católica de Chile. We are grateful to the Administrative Directions of the School of Biological Sciences and the School of Medicine of the Pontificia Universidad Católica de Chile for their administrative support. We would also like to thank to the members of the independent data safety monitoring committee (members in the SA) for their oversight, and finally to the subjects enrolled in the study for their participation and commitment with this trial. This project has been funded in whole or in part with Federal funds from the National Institute of Allergy and Infectious Diseases, National Institutes of Health, Department of Health and Human Services, under Contract No. 75N93021C00016 to A.S. and Contract No. 75N93019C00065 to A.S and D.W.

## Author contributions

**Conceptualization**: AMK, KA, SMB, PAG, JVG, GZ, WM, FM-G, CM, HFP, BMS.

**Visualization:** AMK, KA, SMB, PAG, JVG, GZ, WM.

**Methodology and research:** FM-G, CM, HFP, BMS, AP-I, MR, DM-T, PP-S, DL, CO, CC, NMSG, JAS, LFD, DR-P, YV, AC, SB, CI, MU, MSN, AR, RF, JF, JM, ER, AG-A, MA, FV-E, RS-R, DW, AG, AS, GZ, WM, JVG-A

**Data analysis:** FM-G, CM, HFP, BMS, AP-I, MR, DM-T, LFD, AG-A, MA, FV-E, RS-R

**Funding acquisition:** AMK, SMB.

**Project administration:** AMK, KA, SMB, PAG.

**Supervision:** AMK, KA, SMB, PAG.

**Writing – original draft:** FM-G, CM, HFP, BMS.

**Writing – review & editing:** AMK, SMB, PAG, FM-G, CM, HFP, BMS.

**Verifying underlying data:** AMK, SMB, FM-G, CM, HFP, BMS.

Members of the CoronaVac03CL study group are listed in the supplementary material

## Notes

### Clinical Trial

NCT04651790

### Clinical Protocols

https://clinicaltrials.gov/ct2/show/NCT04651790

### Author Declarations

Samples analyzed in this study were obtained from volunteers recruited in the clinical trial CoronaVac03CL (clinicaltrials.gov #NCT04651790) in Chile (November 2020 to current date). Moreover, the study protocol was reviewed and approved by the Institutional Scientific Ethical Committee of Health Sciences at the Pontificia Universidad Catolica de Chile (#200708006) and the trial was approved by the Chilean Public Health Institute (#24204/20) and conducted according to the current Tripartite Guidelines for Good Clinical Practices, the Declaration of Helsinki, and local regulations. Informed consent was obtained from all volunteers upon enrollment.

### Summary of Updates

Table 1 was updated after a more rigorous statistical analysis. Respective paragraphs in the result, discussion and method sections were accordingly updated.

## References

1. Planas, D., Saunders, N., Maes, P., Guivel-Benhassine, F., Planchais, C., Buchrieser, J., Bolland, W.H., Porrot, F., Staropoli, I., Lemoine, F., et al. (2022). Considerable escape of SARS-CoV-2 Omicron to antibody neutralization. Nature 602, 671–675. 10.1038/s41586-021-04389-z.

2. Levin, E.G., Lustig, Y., Cohen, C., Fluss, R., Indenbaum, V., Amit, S., Doolman, R., Asraf, K., Mendelson, E., Ziv, A., et al. (2021). Waning Immune Humoral Response to BNT162b2 Covid-19 Vaccine over 6 Months. N Engl J Med 385, e84. 10.1056/NEJMoa2114583.

3. Nagy, A., and Alhatlani, B. (2021). An overview of current COVID-19 vaccine platforms. Comput Struct Biotechnol J 19, 2508–2517. 10.1016/j.csbj.2021.04.061.

4. Magen, O., Waxman, J.G., Makov-Assif, M., Vered, R., Dicker, D., Hernan, M.A., Lipsitch, M., Reis, B.Y., Balicer, R.D., and Dagan, N. (2022). Fourth Dose of BNT162b2 mRNA Covid-19 Vaccine in a Nationwide Setting. N Engl J Med 386, 1603–1614. 10.1056/NEJMoa2201688.

5. Reyes, H., Diethelm-Varela, B., Mendez, C., Rebolledo-Zelada, D., Lillo-Dapremont, B., Munoz, S.R., Bueno, S.M., Gonzalez, P.A., and Kalergis, A.M. (2022). Contribution of Two-Dose Vaccination Toward the Reduction of COVID-19 Cases, ICU Hospitalizations and Deaths in Chile Assessed Through Explanatory Generalized Additive Models for Location, Scale, and Shape. Front Public Health 10, 815036. 10.3389/fpubh.2022.815036.

6. Gao, Q., Bao, L., Mao, H., Wang, L., Xu, K., Yang, M., Li, Y., Zhu, L., Wang, N., Lv, Z., et al. (2020). Development of an inactivated vaccine candidate for SARS-CoV-2. Science 369, 77–81. 10.1126/science.abc1932.

7. Chen, Z., Zheng, W., Wu, Q., Chen, X., Peng, C., Tian, Y., Sun, R., Dong, J., Wang, M., Zhou, X., et al. (2022). Global diversity of policy, coverage, and demand of COVID-19 vaccines: a descriptive study. BMC Med 20, 130. 10.1186/s12916-022-02333-0.

8. Tanriover, M.D., Doganay, H.L., Akova, M., Guner, H.R., Azap, A., Akhan, S., Kose, S., Erdinc, F.S., Akalin, E.H., Tabak, O.F., et al. (2021). Efficacy and safety of an inactivated whole-virion SARS-CoV-2 vaccine (CoronaVac): interim results of a double-blind, randomised, placebo-controlled, phase 3 trial in Turkey. Lancet 398, 213–222. 10.1016/S0140-6736(21)01429-X.

9. Han, B., Song, Y., Li, C., Yang, W., Ma, Q., Jiang, Z., Li, M., Lian, X., Jiao, W., Wang, L., et al. (2021). Safety, tolerability, and immunogenicity of an inactivated SARS-CoV-2 vaccine (CoronaVac) in healthy children and adolescents: a double-blind, randomised, controlled, phase 1/2 clinical trial. Lancet Infect Dis 21, 1645–1653. 10.1016/S1473-3099(21)00319-4.

10. Seyahi, E., Bakhdiyarli, G., Oztas, M., Kuskucu, M.A., Tok, Y., Sut, N., Ozcifci, G., Ozcaglayan, A., Balkan, II, Saltoglu, N., et al. (2021). Antibody response to inactivated COVID-19 vaccine (CoronaVac) in immune-mediated diseases: a controlled study among hospital workers and elderly. Rheumatol Int 41, 1429–1440. 10.1007/s00296-021-04910-7.

11. Jara, A., Undurraga, E.A., Gonzalez, C., Paredes, F., Fontecilla, T., Jara, G., Pizarro, A., Acevedo, J., Leo, K., Leon, F., et al. (2021). Effectiveness of an Inactivated SARS-CoV-2 Vaccine in Chile. N Engl J Med 385, 875–884. 10.1056/NEJMoa2107715.

12. Bueno, S.M., Abarca, K., Gonzalez, P.A., Galvez, N.M.S., Soto, J.A., Duarte, L.F., Schultz, B.M., Pacheco, G.A., Gonzalez, L.A., Vazquez, Y., et al. (2021). Safety and Immunogenicity of an Inactivated SARS-CoV-2 Vaccine in a Subgroup of Healthy Adults in Chile. Clin Infect Dis. 10.1093/cid/ciab823.

13. Melo-Gonzalez, F., Soto, J.A., Gonzalez, L.A., Fernandez, J., Duarte, L.F., Schultz, B.M., Galvez, N.M.S., Pacheco, G.A., Rios, M., Vazquez, Y., et al. (2021). Recognition of Variants of Concern by Antibodies and T Cells Induced by a SARS-CoV-2 Inactivated Vaccine. Front Immunol 12, 747830. 10.3389/fimmu.2021.747830.

14. Falsey, A.R., Frenck, R.W., Jr., Walsh, E.E., Kitchin, N., Absalon, J., Gurtman, A., Lockhart, S., Bailey, R., Swanson, K.A., Xu, X., et al. (2021). SARS-CoV-2 Neutralization with BNT162b2 Vaccine Dose 3. N Engl J Med 385, 1627–1629. 10.1056/NEJMc2113468.

15. Choi, A., Koch, M., Wu, K., Chu, L., Ma, L., Hill, A., Nunna, N., Huang, W., Oestreicher, J., Colpitts, T., et al. (2021). Safety and immunogenicity of SARS-CoV-2 variant mRNA vaccine boosters in healthy adults: an interim analysis. Nat Med 27, 2025–2031. 10.1038/s41591-021-01527-y.

16. Pegu, A., O’Connell, S.E., Schmidt, S.D., O’Dell, S., Talana, C.A., Lai, L., Albert, J., Anderson, E., Bennett, H., Corbett, K.S., et al. (2021). Durability of mRNA-1273 vaccine-induced antibodies against SARS-CoV-2 variants. Science 373, 1372–1377. 10.1126/science.abj4176.

17. Moreira, E.D., Jr., Kitchin, N., Xu, X., Dychter, S.S., Lockhart, S., Gurtman, A., Perez, J.L., Zerbini, C., Dever, M.E., Jennings, T.W., et al. (2022). Safety and Efficacy of a Third Dose of BNT162b2 Covid-19 Vaccine. N Engl J Med 386, 1910–1921. 10.1056/NEJMoa2200674.

18. Schultz, B.M., Melo-Gonzalez, F., Duarte, L.F., Galvez, N.M.S., Pacheco, G.A., Soto, J.A., Berrios-Rojas, R.V., Gonzalez, L.A., Moreno-Tapia, D., Rivera-Perez, D., et al. (2022). A Booster Dose of CoronaVac Increases Neutralizing Antibodies and T Cells that Recognize Delta and Omicron Variants of Concern. mBio, e0142322. 10.1128/mbio.01423-22.

19. Regev-Yochay, G., Gonen, T., Gilboa, M., Mandelboim, M., Indenbaum, V., Amit, S., Meltzer, L., Asraf, K., Cohen, C., Fluss, R., et al. (2022). Efficacy of a Fourth Dose of Covid-19 mRNA Vaccine against Omicron. N Engl J Med 386, 1377–1380. 10.1056/NEJMc2202542.

20. Duarte, L.F., Galvez, N.M.S., Iturriaga, C., Melo-Gonzalez, F., Soto, J.A., Schultz, B.M., Urzua, M., Gonzalez, L.A., Vazquez, Y., Rios, M., et al. (2021). Immune Profile and Clinical Outcome of Breakthrough Cases After Vaccination With an Inactivated SARS-CoV-2 Vaccine. Front Immunol 12, 742914. 10.3389/fimmu.2021.742914.

21. Abarca, K., Iturriaga, C., Urzua, M., Le Corre, N., Pineda, A., Fernandez, C., Dominguez, A., Gonzalez, P.A., Bueno, S.M., Donato, P., et al. (2022). Safety and Non-Inferiority Evaluation of Two Immunization Schedules with an Inactivated SARS-CoV-2 Vaccine in Adults: A Randomized Clinical Trial. Vaccines (Basel) 10. 10.3390/vaccines10071082.

22. Grifoni, A., Weiskopf, D., Ramirez, S.I., Mateus, J., Dan, J.M., Moderbacher, C.R., Rawlings, S.A., Sutherland, A., Premkumar, L., Jadi, R.S., et al. (2020). Targets of T Cell Responses to SARS-CoV-2 Coronavirus in Humans with COVID-19 Disease and Unexposed Individuals. Cell 181, 1489–1501 e1415. 10.1016/j.cell.2020.05.015.

23. Arbel, R., Sergienko, R., Friger, M., Peretz, A., Beckenstein, T., Yaron, S., Netzer, D., and Hammerman, A. (2022). Effectiveness of a second BNT162b2 booster vaccine against hospitalization and death from COVID-19 in adults aged over 60 years. Nat Med 28, 1486–1490. 10.1038/s41591-022-01832-0.

24. Tuekprakhon, A., Nutalai, R., Dijokaite-Guraliuc, A., Zhou, D., Ginn, H.M., Selvaraj, M., Liu, C., Mentzer, A.J., Supasa, P., Duyvesteyn, H.M.E., et al. (2022). Antibody escape of SARS-CoV-2 Omicron BA.4 and BA.5 from vaccine and BA.1 serum. Cell 185, 2422–2433 e2413. 10.1016/j.cell.2022.06.005.

25. Hurme, A., Jalkanen, P., Heroum, J., Liedes, O., Vara, S., Melin, M., Terasjarvi, J., He, Q., Poysti, S., Hanninen, A., et al. (2022). Long-Lasting T Cell Responses in BNT162b2 COVID-19 mRNA Vaccinees and COVID-19 Convalescent Patients. Front Immunol 13, 869990. 10.3389/fimmu.2022.869990.

26. Tarke, A., Coelho, C.H., Zhang, Z., Dan, J.M., Yu, E.D., Methot, N., Bloom, N.I., Goodwin, B., Phillips, E., Mallal, S., et al. (2022). SARS-CoV-2 vaccination induces immunological T cell memory able to cross-recognize variants from Alpha to Omicron. Cell 185, 847–859 e811. 10.1016/j.cell.2022.01.015.

27. Zhang, Z., Mateus, J., Coelho, C.H., Dan, J.M., Moderbacher, C.R., Galvez, R.I., Cortes, F.H., Grifoni, A., Tarke, A., Chang, J., et al. (2022). Humoral and cellular immune memory to four COVID-19 vaccines. Cell 185, 2434–2451 e2417. 10.1016/j.cell.2022.05.022.

28. Escobar, A., Reyes-Lopez, F.E., Acevedo, M.L., Alonso-Palomares, L., Valiente-Echeverria, F., Soto-Rifo, R., Portillo, H., Gatica, J., Flores, I., Nova-Lamperti, E., et al. (2021). Evaluation of the Immune Response Induced by CoronaVac 28-Day Schedule Vaccination in a Healthy Population Group. Front Immunol 12, 766278. 10.3389/fimmu.2021.766278.

29. Mok, C.K.P., Cohen, C.A., Cheng, S.M.S., Chen, C., Kwok, K.O., Yiu, K., Chan, T.O., Bull, M., Ling, K.C., Dai, Z., et al. (2022). Comparison of the immunogenicity of BNT162b2 and CoronaVac COVID-19 vaccines in Hong Kong. Respirology 27, 301–310. 10.1111/resp.14191.

30. Reynolds, C.J., Swadling, L., Gibbons, J.M., Pade, C., Jensen, M.P., Diniz, M.O., Schmidt, N.M., Butler, D.K., Amin, O.E., Bailey, S.N.L., et al. (2020). Discordant neutralizing antibody and T cell responses in asymptomatic and mild SARS-CoV-2 infection. Sci Immunol 5. 10.1126/sciimmunol.abf3698.

31. Galipeau, Y., Siragam, V., Laroche, G., Marion, E., Greig, M., McGuinty, M., Booth, R.A., Durocher, Y., Cuperlovic-Culf, M., Bennett, S.A.L., et al. (2021). Relative Ratios of Human Seasonal Coronavirus Antibodies Predict the Efficiency of Cross-Neutralization of SARS-CoV-2 Spike Binding to ACE2. EBioMedicine 74, 103700. 10.1016/j.ebiom.2021.103700.

32. Beltran-Pavez, C., Riquelme-Barrios, S., Oyarzun-Arrau, A., Gaete-Argel, A., Gonzalez-Stegmaier, R., Cereceda-Solis, K., Aguirre, A., Travisany, D., Palma-Vejares, R., Barriga, G.P., et al. (2021). Insights into neutralizing antibody responses in individuals exposed to SARS-CoV-2 in Chile. Sci Adv 7. 10.1126/sciadv.abe6855.

33. Zhang, Y., Zeng, G., Pan, H., Li, C., Hu, Y., Chu, K., Han, W., Chen, Z., Tang, R., Yin, W., et al. (2021). Safety, tolerability, and immunogenicity of an inactivated SARS-CoV-2 vaccine in healthy adults aged 18-59 years: a randomised, double-blind, placebo-controlled, phase 1/2 clinical trial. Lancet Infect Dis 21, 181–192. 10.1016/S1473-3099(20)30843-4.

34. Dan, J.M., Mateus, J., Kato, Y., Hastie, K.M., Yu, E.D., Faliti, C.E., Grifoni, A., Ramirez, S.I., Haupt, S., Frazier, A., et al. (2021). Immunological memory to SARS-CoV-2 assessed for up to 8 months after infection. Science 371. 10.1126/science.abf4063.

