## Supplementary material for "Humoral and cellular response induced by a second booster of an inactivated SARS-CoV-2 vaccine in adults"

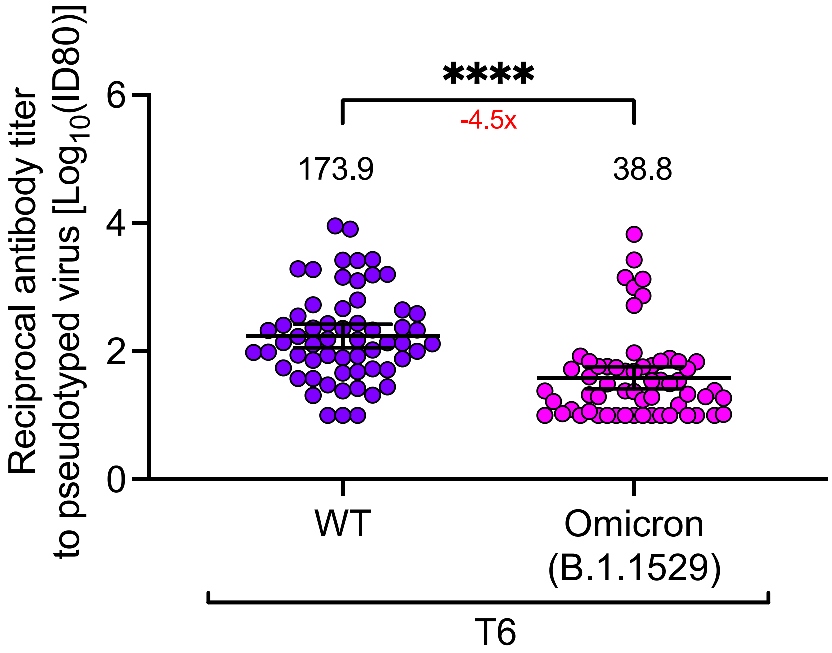


**Supplementary figure 1. Pseudotype-based neutralization assay of SARS-CoV-2.** Neutralizing antibodies were detected in the serum of sixty subjects, four weeks after the second booster dose of CoronaVac®, using a pseudotyped virus neutralization test (pVNT). Data are expressed as the $\log_{10}$ of the reciprocal of the dilution preventing 80% of the infection (ID80). Numbers above each group represent the mean, and the error bars indicate the 95% CI. The number below the significance bar represents the fold decrease of the GMT four weeks after the second booster dose between the WT SARS-CoV-2 and the Omicron variant A parametric t-test was used to compare the neutralization of WT SARS-CoV-2 with the Omicron variant. ****$p<0.0001$

**Supplementary table 1: P values estimated for longitudinal humoral immunity analysis in sera from pre-immune to 4 weeks after the 4th dose.**

| Parameter evaluated | T1 vs T2 | T1 vs T3 | T1 vs T4 | T1 vs T5 | T1 vs T6 | T2 vs T3 | T3 vs T4 | T4 vs T5 | T5 vs T6 | T4 vs T6 |
|---|---|---|---|---|---|---|---|---|---|---|
| sVNT GMU | <0.0001 | <0.0001 | <0.0001 | <0.0001 | <0.0001 | <0.0001 | <0.0001 | <0.0001 | <0.0001 | >0.9999 |
| cVNT GMT | <0.0001 | <0.0001 | <0.0001 | <0.0001 | <0.0001 | 0.0169 | <0.0001 | 0.6407 | 0.0002 | 0.9361 |

sVNT: Surrogate Virus Neutralization, cVNT: Conventional Virus Neutralization, GMU: Geometric mean units, GMT: Geometric mean titer

**Supplementary table 2: P values estimated for humoral and cellular immunity against the Delta and the Omicron variants 4 weeks after the 3rd dose and 4 weeks after the 4th dose.**

| Parameter evaluated | T4 vs T6 WT | T4 vs T6 Delta | T4 vs T6 Omicron | T4 WT vs T4 Omicron | T6 WT vs T6 Omicron | T4 WT vs T4 Delta | T6 WT vs T6 Delta |
|---|---|---|---|---|---|---|---|
| sVNT GMT | 0.0016 | >0.9999 | 0.0087 | <0.0001 | <0.0001 | <0.0001 | 0.0069 |
| AIM+($OX40^+CD137^+$) | 0.6396 | 0.2714 | 0.6747 | 0.9819 | 0.9748 | 0.9983 | 0.9103 |
| IFN-$\gamma^+$ SFC MP-S variants | 0.6987 | >0.9999 | >0.9999 | >0.9999 | >0.9999 | 0.7426 | 0.9712 |

sVNT: Surrogate Virus Neutralization; GMT: Geometric mean titer; AIM: Activation-Induced-Marker; SFC: Spot Forming Cells; MP-S: Spike mega-pool of peptides

**Supplementary table 3: Seropositivity rates, seroconversion rates, and Geometric Mean Titers (GMT) and Units (GMU) of circulating neutralizing antibodies against SARS-CoV-2 RBD*.**

| Methodology** | Indicators | T2 | T3 | T4 | T5 | T6 |
|---|---|---|---|---|---|---|
| sVNT | Seropositivity | 81/97 | 63/87 | 84/87 | 79/87 | 84/87 |
| | % | 93.1 | 72.4 | 96.6 | 90.8 | 96.6 |
| | Seroconversion | 70/87 | 32/87 | 81/87 | 63/87 | 81/87 |
| | % | 80.5 | 36.8 | 93.1 | 72.4 | 93.1 |
| | GMU | 199.7 | 53.1 | 586.0 | 220.4 | 549.2 |
| | 95% CI | 154.9-257.4 | 43.3-65.2 | 447.8-766.7 | 154.9-313.4 | 418.2-721.2 |
| cVNT | Seropositivity | 52/63 | 42/63 | 62/63 | 57/63 | 62/63 |
| | % | 82.5 | 66.7 | 98.4 | 90.5 | 98.4 |
| | Seroconversion | 37/63 | 18/63 | 54/63 | 45/63 | 60/63 |
| | % | 58.7 | 28.6 | 85.7 | 71.4 | 95.2 |
| | GMT | 21.1 | 9.9 | 95.1 | 54.9 | 149.2 |
| | 95% CI | 13.9-21.8 | 6.8-14.6 | 62.7-144.3 | 32.3-90.3 | 94.6-235.8 |

*Samples from volunteers were used to evaluate the neutralizing capacity of antibodies at the different times evaluated. 87 samples were evaluated by sVNT assays and 63 by cVNT assays.
**sVNT: Surrogate Virus Neutralization; cVNT: Conventional Virus Neutralization; GMU: Geometric mean units; GMT: Geometric mean titer; BDM: Receptor Binding Domain

**Supplementary table 4: Seropositivity rates, seroconversion rates and Geometric Mean Titers (GMT) of circulating neutralizing antibodies against SARS-CoV-2 RBD of Delta and Omicron variant.**

| Methodology | Indicators | T4 | T6 |
|---|---|---|---|
| Delta (B.1.617.2) | Seropositivity | 81/87 | 82/87 |
| | % | 93.1 | 94.3 |
| | Seroconversion | 70/87 | 59/87 |
| | % | 80.5 | 67.8 |
| | GMT | 21.5 | 21.8 |
| | 95% CI | 17.2-26.8 | 16.8-28.4 |
| Omicron (B.1.1.529) | Seropositivity | 15/87 | 28/87 |
| | % | 17.2 | 32.2 |
| | Seroconversion | 4/87 | 16/87 |
| | % | 4.6 | 18.4 |
| | GMT | 2.5 | 3.7 |
| | 95% CI | 2.2-2.8 | 2.9-4.7 |

GMT: Geometric mean titer

**Supplementary table 5: P values estimated for longitudinal cellular immunity assays from pre-immune to 4 weeks after the 4th dose.**

| Parameter evaluated | T1 vs T2 | T1 vs T3 | T1 vs T4 | T1 vs T5 | T1 vs T6 | T2 vs T3 | T3 vs T4 | T4 vs T5 | T5 vs T6 | T4 vs T6 |
|---|---|---|---|---|---|---|---|---|---|---|
| CD4+AIM+(OX40$^+$CD13) | 0.0241 | <0.0001 | <0.0001 | 0.0041 | 0.0684 | 0.2360 | 0.6536 | 0.0284 | 0.9984 | 0.6560 |
| CD8+AIM+(CD69$^+$CD13) | >0.9999 | >0.9999 | >0.9999 | >0.9999 | >0.9999 | >0.9999 | >0.9999 | >0.9999 | >0.9999 | >0.9999 |
| IFN-γ$^+$ SFC MP-S+R | 0.0004 | 0.0037 | <0.0001 | 0.0009 | <0.0006 | >0.9999 | 0.8717 | 0.9108 | >0.9999 | 0.9148 |
| IFN-γ$^+$ SFC MP-CD8A+B | 0.6047 | >0.9999 | 0.1957 | 0.0546 | 0.2362 | >0.9999 | 0.9595 | 0.9984 | >0.9999 | 0.9998 |

AIM: Activation-Induced-Marker; SFC: Spot Forming Cells; MP-S: Spike mega-pool of peptides

**Supplementary table 6: Frequency of volunteers with detectable T cell response against the Spike protein of SARS-CoV-2 WT, Delta and the Omicron variants.**

| SARS-CoV-2 variant | Methodology | Indicators | T4 | T6 |
|---|---|---|---|---|
| WT | Flow Cytometry | CD4+AIM+(OX40$^+$CD13$^+$) | 26/46 | 15/46 |
| | | % | 56.5 | 32.6 |
| | ELISPOT | IFN-γ$^+$ SFC MP-S | 25/46 | 25/46 |
| | | % | 54.3 | 54.3 |
| Delta (B.1.617.2) | Flow Cytometry | CD4+AIM+(OX40$^+$CD13$^+$) | 25/46 | 9/46 |
| | | % | 54.3 | 19.6 |
| | ELISPOT | IFN-γ$^+$ SFC MP-S | 28/46 | 29/46 |
| | | % | 60.9 | 63.0 |
| Omicron (B.1.1.529) | Flow Cytometry | CD4+AIM+(OX40$^+$CD13$^+$) | 29/46 | 14/46 |
| | | % | 63.0 | 30.4 |
| | ELISPOT | IFN-γ$^+$ SFC MP-S | 26/46 | 21/46 |
| | | % | 56.5 | 45.6 |

**Supplementary table 7: List of antibodies for flow cytometry**

| Marker | Clone | Fluorophore | Supplier | Dilution |
|---|---|---|---|---|
| CD3 | OKT3 | Alexa Fluor 700 | BioLegend | 1:50 |
| CD4 | RPA-T4 | BV605 | BioLegend | 1:50 |
| CD8 | RPA-T8 | BV650 | BioLegend | 1:50 |
| CD14 | M5E2 | V500 | BD | 1:100 |
| CD19 | HIB19 | V500 | BD | 1:100 |
| CD69 | FN50 | PE | BD | 1:10 |
| CD137 | 4-1BB | APC | BioLegend | 1:50 |
| OX40 | BER-ACT35 | PE-Cy7 | BioLegend | 1:50 |
| Fixable Viability Dye | - | BV510 | BD | 1:1000 |

1. CoronaVac03CL Study Group

Center CL1: Áreas Ambulatorias Marcoleta - Pontificia Universidad Católica de Chile.
Álvaro Rojas, María Soledad Navarrete, Constanza Del Río, Dinely Del Pino, Natalia Aguirre, Grecia Salinas, Franco Vega, Acsa Salgado, Thomas Quinteros, Marlene Ortiz, Marcela Puente, Alma Muñoz, Patricio Astudillo, Nicole Le Corre.

Center CL2: Clínica San Carlos de Apoquindo - Red de Salud UC-Christus
Marcela Potin, Juan Catalán, Melan Peralta, Consuelo Zamanillo, Nicole Keller, Rocío Fernández, Sofía Aljaro, Sofía López, José Tomás González, Tania Weil, Luz Opazo, Paula Muñoz, Inés Estay, Miguel Cantillana, Liliana Carrera, Matías Masalleras.

Center CL4: Clínica Los Andes – Universidad de Los Andes
Paula Guzmán, Francisca Aguirre, Aarón Cortés, Luis Federico Bátiz, Javiera Pérez, Karen Apablaza, Lorena Yates, María de los Ángeles Valdés, Bernardita Hurtado, Veronique Venteneul, Constanza Astorga.

Center CL5: Clínica Alemana - Universidad del Desarrollo
Paula Muñoz-Venturelli, Pablo A. Vial, Andrea Schilling, Daniela Pavez, Inia Pérez, Amy Riviotta, Francisca González, Francisca Urrutia, Alejandra Del Río, Claudia Asenjo, Bárbara Vargas, Francisca Castro, Alejandra Acuña, Javiera Guzmán, Camila Astudillo.

Center CL6: Hospital Clínico Félix Bulnes - Universidad San Sebastián
Carlos M. Pérez, Pilar Espinoza, Andrea Martínez, Marcela Arancibia, Harold Romero, Cecilia Bustamante, María Loreto Pérez, Natalia Uribe, Viviana Silva, Bernardita Morice, Marco Pérez.

Center CL7: Hospital Dr. Gustavo Fricke - Universidad de Valparaíso

Marcela González, Werner Jensen, Claudia Pasten, M. Fernanda Aguilera, Nataly Martínez, Camila Molina, Sebastián Arrieta, Begoña López, Claudia Ortiz, Macarena Escobar, Camila Bustamante, Marcia Espinoza, Angela Pardo, Alison Carrasco, Miguel Montes, Macarena Saldías, Natalia Gutiérrez, Juliette Sánchez.

Center CL8: Hospital Carlos Van Buren- Universidad de Valparaíso
Daniela Fuentes, Yolanda Calvo, Mariela Cepeda, Rosario Lemus, Muriel Suárez, Mercedes Armijo, Shirley Monsalves, Constance Marucich, Cecilia Cornejo, Ángela Acosta, Xaviera Prado, Francisca Yáñez, Marisol Barroeta, Claudia López.

Center CL9: Complejo Asistencial Dr. Sótero del Rio
Paulina Donato, Martin Lasso, María Iturrieta, Juan Giraldo, Francisco Gutiérrez, María Acuña, Ada Cascone, Raymundo Rojas, Camila Sepúlveda, Mario Contreras,

Yessica Campisto, Pablo González, Zoila Quizhpi, Mariella López, Vania Pizzeghello, Stephannie Silva.

**2. Members of the Independent Data Safety Monitoring Committee.**

Luis Delpiano, MD, Pediatric Infectious Diseases Specialist, Hospital San Borja Arriarán, Santiago, Chile.
Gloria Icaza, MD, Epidemiologist and Statistician, Universidad de Talca, Talca, Chile. Leonardo Chanqueo, MD, Infectious Disaeses Specialist, Hospital San Juan de Dios, Santiago, Chile.
Mónica Imarai, PhD, Universidad de Santiago, Santiago, Chile.
